# Current status and future opportunities in modeling Multiple Sclerosis clinical characteristics

**DOI:** 10.1101/2022.02.24.22271474

**Authors:** Joshua Liu, Erin Kelly, Bibiana Bielekova

## Abstract

Development of effective treatments requires understanding of disease mechanisms. For diseases of the central nervous system (CNS), like Multiple sclerosis (MS), human pathology studies and animal models tend to identify candidate disease mechanisms. However, these studies cannot easily link identified processes to clinical outcomes, such as MS severity, required for causality assessment of candidate mechanisms. Technological advances now allow generation of thousands of biomarkers in living human subjects, derived from genes, transcripts, medical images and proteins or metabolites in biological fluids. These biomarkers can be assembled into computational models of clinical value, provided such models are generalizable. Reproducibility of models increases with technical rigor of study design, such as blinding, implementing controls, using large cohorts that encompass entire spectrum of disease phenotypes and, most importantly, validating models in independent cohort(s).

To facilitate growth of this important research area, we performed a meta-analysis of publications that model MS clinical outcomes (n=302), extracting effect sizes, while also scoring technical quality of study design using pre-defined criteria. Finally, we generated a Shiny-App-based website that allows dynamic exploration of data using selective filtering.

On average, published studies fulfilled only one out of seven criteria of study design rigor. Only 15.2% of studies used any validation strategy, and only 8% used the gold standard of independent cohort validation. Many studies also used small cohorts, e.g., for MRI and blood biomarker predictors the median sample size was below 100 subjects. We observed inverse relationships between reported effect sizes and the numbers of study design criteria fulfilled, expanding analogous reports from non-MS fields, that studies that fail to limit bias over-estimate effect sizes.

In conclusion, the presented meta-analysis represents a useful tool for researchers, reviewers, and funders to improve design of future modeling studies in MS and to easily compare new studies with published literature. We expect that this will accelerate research in this important area, leading to development of robust models with proven clinical value.

## 1 Introduction

Multiple sclerosis (MS) is polygenic, immune-mediated, demyelinating disease of the CNS that causes substantial personal and societal burden. Understanding the pathophysiology of initial stages of MS revealed that focal influx of immune cells to CNS tissue can be non-invasively monitored by contrast enhancing lesions (CELs) on brain MRI [1]. CELs, as surrogate of focal inflammation, allowed rapid screening of therapeutic agents [2], identifying many treatments that effectively block formation of MS lesions.

However, these treatments are not curative, and their efficacy decreases with the advancing age of treatment initiation. Indeed, after age of approximately 54 years no net benefit on disability progression can be demonstrated in Phase III clinical trials [3]. This is partially due to inflammation becoming compartmentalized to CNS tissue during MS evolution [4, 5], making it largely inaccessible to systemically administered treatments. But neurodegenerative mechanisms [6, 7] likely contribute to the decreasing efficacy of immunomodulatory treatments. To develop effective treatments of MS beyond inhibiting formation of focal lesions, the MS field must expand its earlier success in gaining pathophysiological insight from early to late disease mechanisms.

Therefore, future therapeutic progress in MS requires identification and validation of biomarkers that reflect mechanisms that cause development of clinical disability at later stages of MS or in patients who no longer form MS lesions thanks to current immunomodulatory treatments. Due to the complexity of these later pathophysiological mechanisms, it is unlikely that a single biomarker can replicate the success of CELs. Indeed, an ability of a single biomarker to reflect key patient-specific outcomes, namely clinical disability and the rate of its development (measured by MS severity outcomes [8]) is extremely limited. Consequently, investigators use simple or complex statistical techniques (including machine learning [ML]) to aggregate biomarkers into models with enhanced predictive power.

To our best knowledge there is no review that summarizes state-of-the-art modeling strategies in MS. The goal of this paper is to present such critical meta-analysis, to help the MS community, including funders, to identify gaps and opportunities in this important research. We performed systematic assessment of the technical quality of reviewed studies, such as sample size, blinding, adjustment for covariates, adjustment for multiple comparisons, integration of healthy volunteer data to differentiate physiological processes such as aging and gender effects from MS-driven pathologies and, most importantly, we evaluated the level of model’s validation. Because it has been repeatedly demonstrated that low technical quality [9, 10] and small sample sizes [11-13] over-estimate effect sizes and lower the probability that the results are reproducible [14, 15], the attributes we summarized are essential determinants of the generalizability of published models. The wide domain of knowledge included in this work can be utilized as a reference for MS researchers, funders, and reviewers.

## 2 Methods

### 2.1 Search Method

We conducted a literature search to identify studies that generated statistical models to predict clinical outcomes among MS patients. This systematic review was conducted in accordance with the Preferred Reporting Items for Systematic Reviews and Meta-Analyses (PRISMA) guidelines. PubMed searches were performed using keywords relating to multiple sclerosis, predictive models, and outcomes. Five PubMed searches were performed to identify relevant MRI studies, using various combinations of the following keywords: “multiple sclerosis,” “disability,” “correlate,” “MRI,” “machine learning,” “predict,” “AI,” “artificial intelligence,” and “neuroimaging.” Two searches were performed to identify other relevant studies reporting statistical modeling in MS with the following PubMed search criteria: “((Multiple Sclerosis[Title/Abstract]) AND (Prediction) AND (Outcome) AND (Model OR Machine Learning)),” on 5/24/2021 and “(((Multiple Sclerosis[Title/Abstract]) AND (Prediction[Title/Abstract]) AND (Outcome))” on 8/16/2021.

### 2.2 Exclusion Criteria

Two reviewers (J.L. and E.K.) independently screened studies that reported effect sizes for image-, clinical-, or biomarker-based models predicting a clinical outcome. We excluded studies with no predictive models, studies with no imaging, clinical, or biomarker predictors, studies with no clinical outcomes, non-human studies, non-MS studies, and studies where full text was not available.

### 2.3 Information Extraction

The following features were extracted from the methods and results of these studies: 1. types of predictors used for the modeling (i.e., clinical, MRI, blood biomarkers, CSF biomarkers, genes); 2. clinical outcome(s) modeled (e.g. EDSS, SPMS Conversion); 3. cohort sample size; 4. all reported effect sizes (e.g. for modeling continuous outcomes: R^2 [i.e., coefficient of determination; a statistical measure of how well the regression prediction approximate the measured data], Spearman Rho [a non-parametric correlation coefficient that measures the strength of association between two variables], Pearson R [a parametric correlation coefficient that measures the strength of association between two variables; should be used only with normally distributed data as it is very sensitive to the effect of outliers]; for dichotomized outcomes such as progression or non-progression: Hazard Ratios [HR: i.e., an estimate of the ratio of the hazard rate such as disability progression in one versus other groups: e.g., in treated versus untreated patients], Odds Ratios [OR; i.e., the cumulative measure of association between events A and B; with OR=1 signifying independence between A and B, while OR>1 signify that A and B are positively associated while OR<1 means that A and B are negatively associated] and finally, p-value [i.e., the probability of obtaining results at least as extreme as observed if the null hypothesis was correct; please note that because p-value depends not only on effect size but also on variance and cohort size, it is extremely poor indicator of effect size alone].

We also extracted seven dichotomized/categorical factors used to assess quality of the study design (see below). We’ll refer to these as indicators of “technical quality” of the study.

### 2.4 Assessment of the quality of study design in the reviewed models

Following seven indicators of technical quality were extracted from the methods and results sections of each paper: 1. Presence and type of model validation (i.e., A. independent validation cohort [gold standard] or B. training cohort out-of-bag/cross-validation); 2. Described process of dealing with outliers to prevent bias (yes/no); 3. Described process of dealing with data missingness to prevent bias (yes/no); 4. Blinding (yes/no); 5. Adjusting for covariates (yes/no); 6. The number of comparisons made (i.e., number of predictors multiplied by the number of outcomes) and whether p-values were adjusted for multiple comparisons (yes/no); 7. Controls utilized (yes/no): specifically, healthy controls to differentiate MS disease process from effects of physiological aging or from physiological sexual dimorphism. Depending on how many of these criteria study fulfilled, the quality of study design ranged from 0 to 7.

The Master worksheet containing all these extracted data, as well as PubMedIDentifiers (PMID) of individual papers is provided as Supplementary Table 1.

### 2.5 Validation of published inverse relationships between study design quality and reported effect sizes

Previous studies in non-MS fields showed that 1. Small cohort studies; 2. Studies of low experimental quality and 3. Studies performed in training cohort only, all significantly overestimate true effect sizes [10, 11, 13, 14]. To assess whether the same can be observed in MS field we investigated relationships between the technical quality of studies (including cohort sizes and comparisons of training versus cross-validation versus independent validation cohorts) and the reported effect sizes.

In addition to univariate analyses, we also classified groups of studies based on combination of cohort size and technical quality criteria: studies were considered high quality if they reached 1 standard deviation above the mean for both factors, whereas low quality were 1 standard deviation below the mean for both. To compare all identified low- and high-quality studies (two-sample Wilcoxon [Mann-Whitney] test), we normalized different metrics of effect sizes to yield common metrics ranging from 0-1.

### 2.6 Public Database Exploration Tool

To allow readers independent exploration of data beyond relationships described in this paper, we developed a Shiny app in R version 3.6.1. This application includes selection tools that allow user to select all or specific predictor types, all or specific clinical outcomes and all or specific effect size statistic tools and then generates a set of two-dimensional plots that visualize relationships between extracted features. The user can also rapidly identify PMID for a specific study by clicking a specific point in the two-dimensional plots.

## 3 Results

### 3.1 Clinical Outcomes

663 studies were screened, excluding duplicate records (Figure 1; PRISMA diagram). After applying the exclusion criteria, 302 studies were included in the review. 189 clinical outcomes were predicted across 302 included studies. The breakdown of outcomes by category is shown in Figure 2.

**Figure 1:**
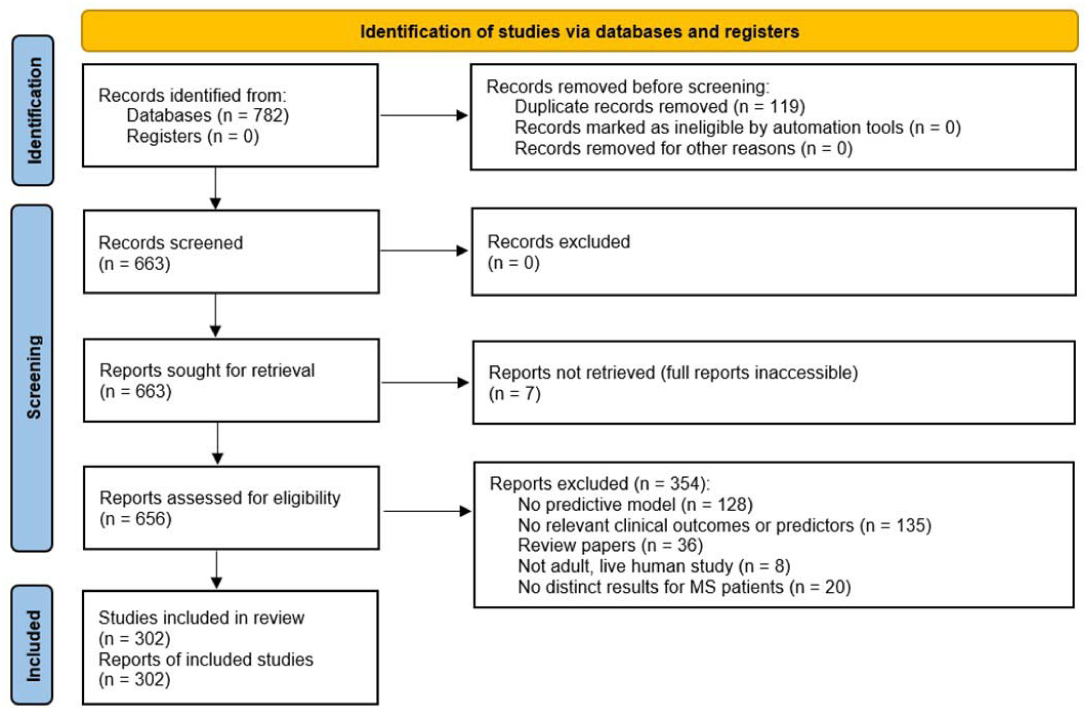
PRISMA chart summarizing the disposition of records identified from PubMed searches. The searches identified 782 records out of which 663 were unique. After various exclusion criteria defined in the figure, 302 unique records were included in the review.

**Figure 2:**
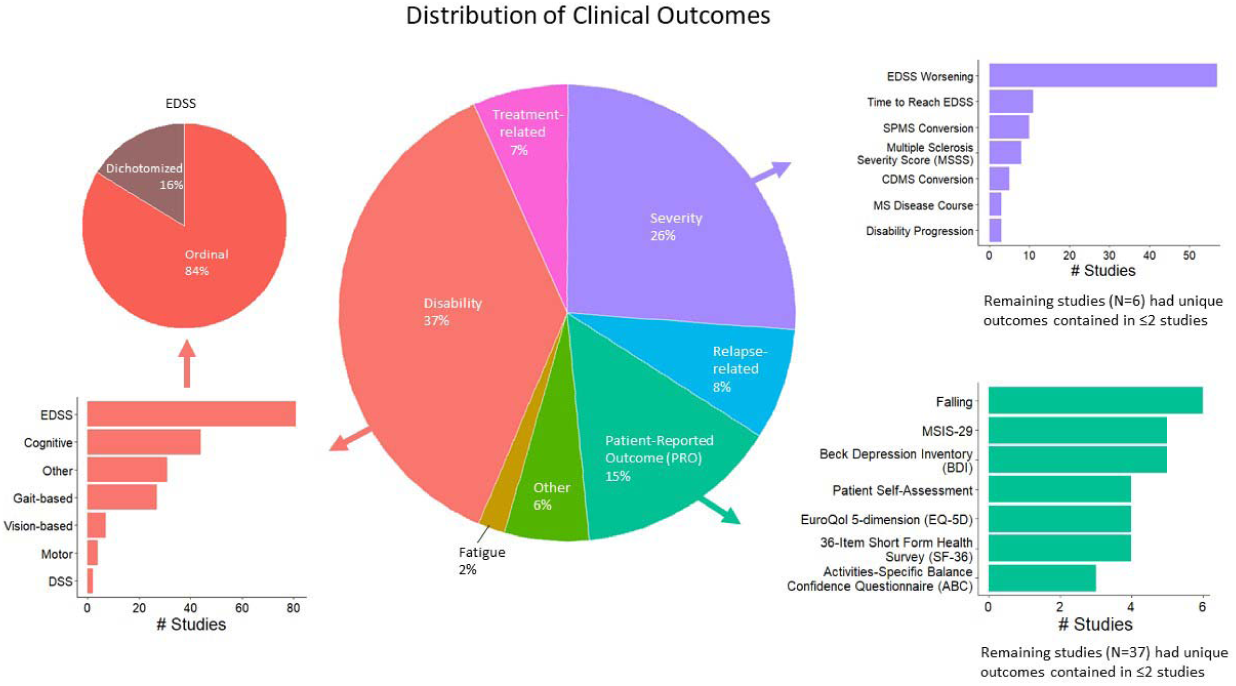
Distribution of modelled clinical outcomes. The large pie chart in the center of the figure shows the distribution of categories of modelled outcomes modeled. The three most frequent outcome categories are disability (37%, red), severity (26%, purple), patient-reported outcomes (15%, teal). The surrounding barplots show the breakdown of each of these three categories.

The largest category of clinical outcomes was *MS progression measured by traditional disability outcomes* (Figure 2, red color; 37% of reviewed studies). Of these, the most prevalent outcomes were EDSS-based (n=81 studies), such as predicting EDSS on an ordinal scale, followed by predicting EDSS as a dichotomous variable. Cognitive disability outcomes constituted second largest sub-category (n=44). These included the Paced Auditory Serial Addition Test [PASAT], the Stroop test, the Symbol Digit Modalities Test [SDMT], and pothers. The third most prevalent progression outcomes were gait based (n=27), which included timed 25-foot walk [T25FW], Hauser ambulation index, 6-minute walk test, Timed Up and Go [TUG], dynamic gait index and others.

Following MS progression/disability outcomes, the next largest category of outcomes was *MS severity outcomes*, which were modelled by 26% of reviewed studies (Figure 2, purple color). 69 studies predicted changes in EDSS over time, including EDSS worsening and time to reach a specified EDSS score. Ten studies predicted conversion to secondary-progressive MS (SPMS), 8 predicted EDSS-based Multiple Sclerosis Severity Score (MSSS), 5 predicted conversions to clinically definite MS and remaining outcomes were studied by less than 5 studies.

Finally, *Patient-Reported Outcomes* (PROs; Figure 2, teal color) were modelled by 15% of reviewed studies. This category was fractionated, with falls predicted inin 6 studies, Multiple Sclerosis Impact Scale (MSIS-29) and Beck Depression Inventory (BDI) by 5 studies each. The remaining outcomes were studied by less than 5 studies.

### 3.2 Predictor Variables

Five categories of predictor variables were used in these models, namely clinical (n=166 studies), MRI (n=103), genes (n=13), blood biomarkers (n=20), and CSF biomarkers (n=9) (Figure 3A).

**Figure 3:**
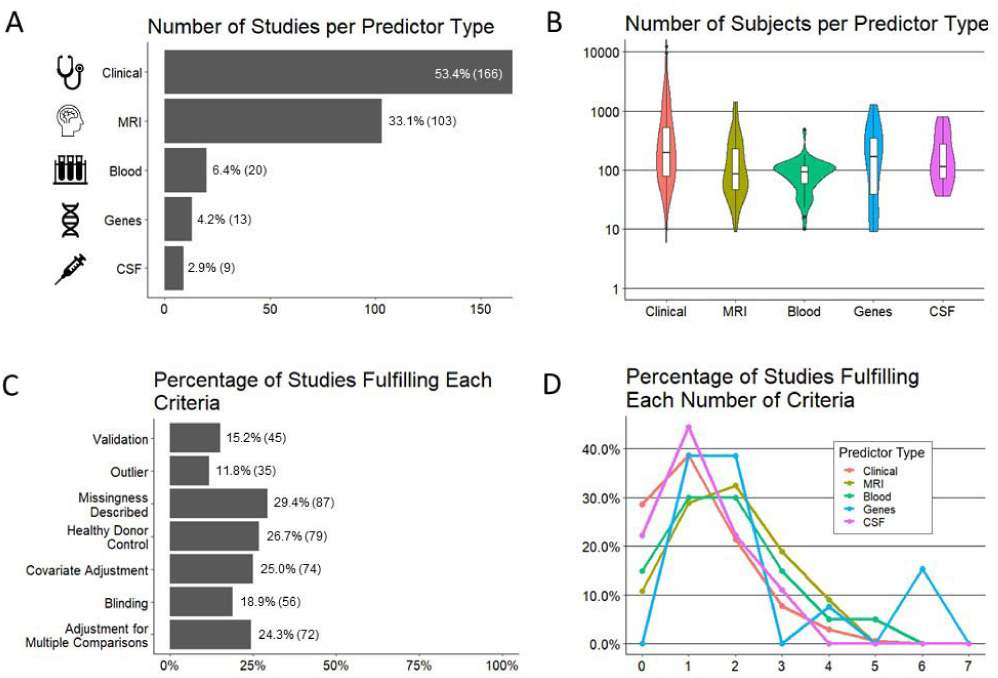
Important characteristics of reviewed studies. (A) number of studies (x-axis) per predictor type (y-axis). (B) number of subjects (y-axis) by predictor type (x-axis). (C) percentage of studies (x-axis; number of studies in the parenthesis) fulfilling each pre-selected criteria of experimental design/technical quality of the study (y-axis). (D) percentage of studies (y-axis) per each predictor type fulfilling a number of the technical criteria (x-axis).

We hypothesized, and confirmed, that the sample sizes would be largest for models using clinical predictors, because they are easiest to collect. Using similar reasoning, we expected smallest sample sizes for CSF predictors due to invasive nature of lumbar punctures. Instead, we observed smallest sample sizes for models utilizing MRI predictors and blood biomarkers, where most studies had sample sizes below 100 patients, with some as low as 10 patients (Figure 3B).

### 3.3 Technical Quality

In addition to recording cohort sizes for each reviewed study, we collected seven factors of study design aimed to minimize bias and therefore maximize probability that the reported results will be generalizable (Figure 3C). These were: 1. Blinded analyses; 2. Pre-defined/described missingness of the data; 3. Pre-defined/described methodology for outlier identification and removal to minimize bias; 4. Adjustment for covariates; 5. Presence of controls, such as healthy volunteers to differentiate physiological processes such as aging or gender effects from MS-related processes; 6. The number of comparisons performed and whether investigators employed any strategy to adjust significance thresholds if the number of comparisons was high; and finally 7. Level of model validation (if any), differentiating cross-validation methods that re-use training cohort samples from true independent cohort validation, considered gold-standard.

Although no study needs to fulfill all 7 criteria to yield reliable results, it was unexpected to observe that majority of studies fulfilled one or less criteria and only 1% of studies fulfilled more than four. Comparing technical quality of studies based on different predictors (Figure 3D), we observed highest technical quality of studies that used genes, followed by MRIs and blood biomarkers. Astonishingly, more than 20% of studies that used clinical or CSF biomarker predictors, fulfilled zero technical quality criteria.

Finally, because current modeling algorithms are highly susceptible to overfit, an essential determinant of model’s generalizability is the level of its validation. Overfit is caused by the ability of ML algorithms to find and amplify subtle changes in the data, including noise, to achieve fit that is much stronger than biologically plausible. Consequently, when the model is applied to new set of samples/patients, it will have much lower fit or may not validate at all. There are two types of validation: First re-uses training cohort data, in various manners that are beyond the scope of this review. It is often called “cross-validation” or “out-of-bag data”. We’ll use term “cross-validation” to signify any validation strategy that reuses training cohort data. To what degree cross-validation faithfully predicts the generalizability of the model depends on the details of how it was performed. Cross-validation may be overly optimistic if researchers failed to prevent bias, and this is often the case. Therefore, the gold standard is independent cohort validation, which implies using model on a new set of samples/subjects that did not contribute, in any way, to model generation.

We observed that only 15% of studies used any type of validation with only 8% of all studies using independent validation.

### 3.4 Effect Sizes

Effect sizes for each of these studies were included as reported (for the explanation of these metrics, see Methods section). The most reported metric was R^2 in 101 studies with Pearson R being reported in 53 studies, hazard ratios in 46 studies, odds ratios in 43 studies, and Spearman Rho in 29 studies. P-values were reported alongside these metrics in 202/302 studies.

Overall, we observed highly selective, rather than comprehensive use of statistical outcomes that reflect effect sizes. This selectivity limits the ability to compare effect sizes between different studies.

### 3.5 Association between study quality and effect size

It is estimated that between 51-89% of published literature in bio-medical sciences is not reproducible [15-17] and poor study design, based on small sample sizes [11, 13] and failure to prevent bias [18-20] is the major contributor to this reproducibility crisis. Indeed, as outlined in the introduction, previous studies highlighted inverse relationship between technical quality of study design [10] (including cohort sizes [11, 13]) and reported effect sizes,, validating the notion that technical quality of study design is major determinant of the generalizability of the gained scientific knowledge.

To assess whether we can identify analogous inverse relationships between reported effect sizes and our pre-defined, systematic grading of technical quality of reported study design, we performed two types of analyses. In first analysis, we compiled all studies that reported any effect size separately for the training (Figure 4A) and cross-validation cohorts (Figure 4B). We then assessed whether there is any relationship between the number of technical quality criteria a study fulfilled versus the reported effect size. For both training cohort data (Figure 4C) and cross-validation (Figure 4D), we observed inverse relationship between the technical quality of the study and the reported effect size.

**Figure 4:**
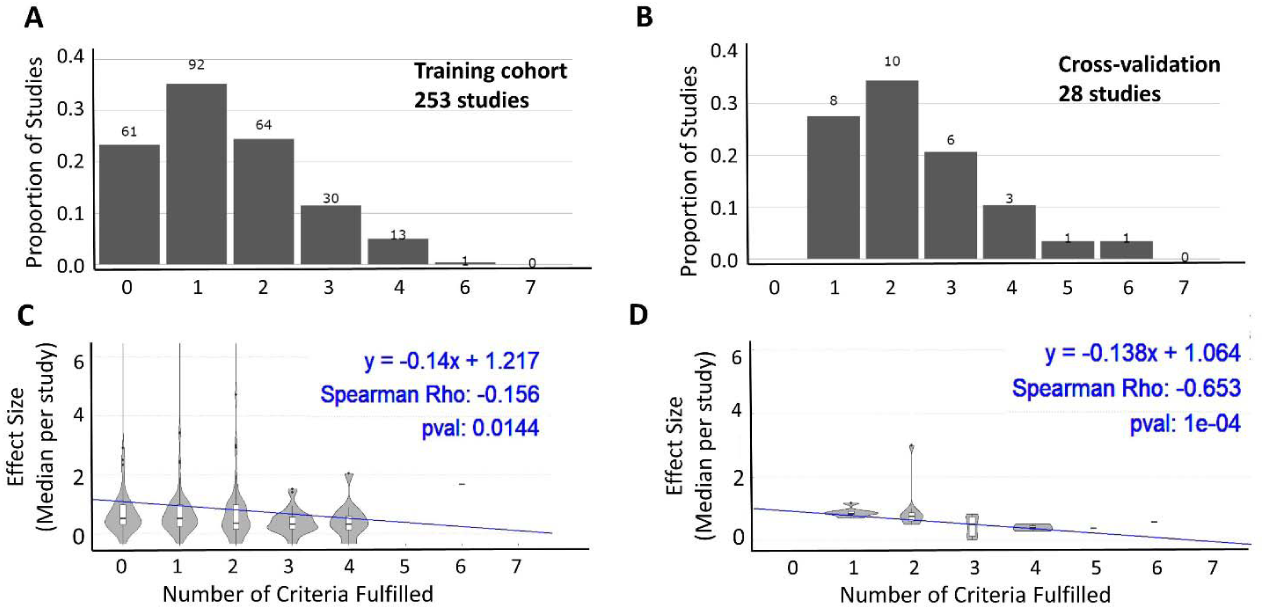
Relationship between technical quality of the study and reported effect size. The proportion of studies fulfilling a sum of the seven technical quality criteria (0 weakest experimental design - 7 strongest experimental design) for 253 studies with training cohorts (A) and cross-validation cohorts (B). The number of studies in each category are listed above the bars. The effect sizes reported by studies categorized based on the number of technical quality criteria they fulfilled (0-7) for training cohort (C; n = 253) and cross-valudation (D; n = 28) results. In both cohorts, the reported effect sizes decreased as number of technical quality criteria these studies fulfilled increased.

Because the above strategy ignored cohort size, which is important determinant of generalizability of the model, in the second analysis we construed two-dimensional assessment of the study design (Figure 5A), integrating both grading of reported technical quality with reported sample sizes. Using means +/- one SD of all studies we identified studies of low quality (i.e., at least 1 SD below the average for both technical quality and sample size) versus high quality (i.e., at least 1 SD above the average for both domains). We observed significantly higher reported standardized effect sizes for the low quality, compared to high quality studies (Figure 5B). As expected, the effect sizes for the remaining studies were centered between the low and high-quality studies.

**Figure 5:**
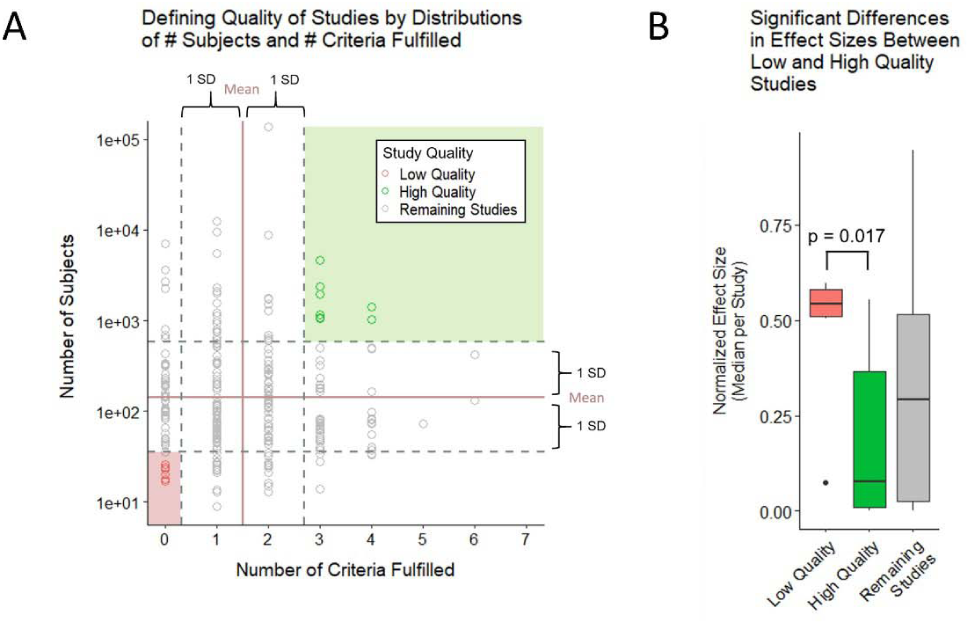
(A) study quality defined by number of subjects and number of criteria fulfilled with high quality studies falling 1 standard deviation above both criteria and low-quality studies falling 1 standard deviation below both criteria. (B) boxplots show comparison between low and high quality effect sizes using a two-sample Wilcoxon (Mann-Whitney) test. Low quality studies were found to have higher effect sizes at a significant p-value of 0.017.

### 3.6 Effect sizes for EDSS-based models of MS progression and MS severity

To facilitate interpretation of any future models, we compared the strength of models for different predictors, using EDSS-based MS progression (Table 1) and MS severity outcomes (MSSS and ARMSS; Table 2). EDSS-based outcomes are the most broadly used in MS field. We found them to be modelled most commonly, and they are accepted by regulatory agencies for assessing therapeutic efficacy of MS drugs. For each outcome and predictor pair, we provide highest reported effect size and the effect size reported by the study of highest technical quality. Whenever available, we also same reported effect sizes for cross-validation and independent validation studies.

**Table 1:**
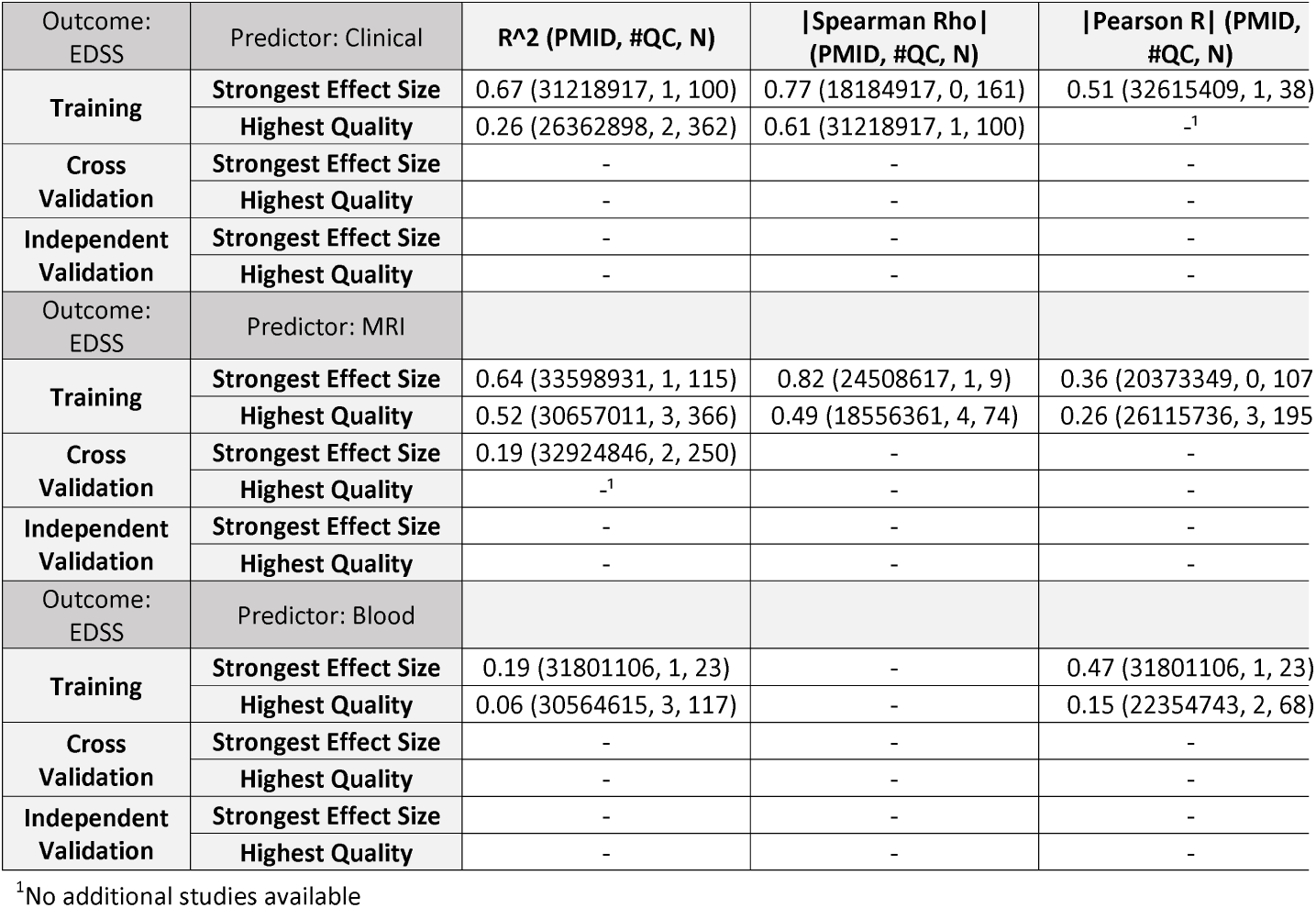
This set of tables shows EDSS outcomes compared to the following predictor types: clinical, MRI, and blood respectively. Effect sizes of studies with the strongest effect sizes were reported as well as for studies with highest quality. The following metrics were explored: R^2, Spearman Rho, and Pearson R.

**Table 2:** This set of tables includes outcomes of MSSS and ARMSS, showing studies with highest effect sizes and highest quality, reporting R^2, Spearman Rho, and Pearson R. Fewer studies looked at these two outcomes, compared to EDSS.

| Outcome:<br>MSSS | Predictor: MRI | R<sup>2</sup> (PMID, #QC, N) | \| Spearman Rho \|<br>(PMID, #QC, N) | \| Pearson R \| (PMID, #<br>N) |
| --- | --- | --- | --- | --- |
| **Training** | **Strongest Effect Size** | 0.45 (24122185, 1, 67) | - | - |
|  | **Highest Quality** | _<sup>1</sup> | - | - |
| **Cross**<br>**Validation** | **Strongest Effect Size** | - | - | - |
|  | **Highest Quality** | - | - | - |
| **Independent**<br>**Validation** | **Strongest Effect Size** | - | - | - |
|  | **Highest Quality** | - | - | - |
| Outcome:<br>MSSS | Predictor: Blood |  |  |  |
| **Training** | **Strongest Effect Size** | 0.24 (20965962, 2, 54) | - | 0.19 (22354743, 2, 6) |
|  | **Highest Quality** | _<sup>1</sup> | - | _<sup>1</sup> |
| **Cross**<br>**Validation** | **Strongest Effect Size** | - | - | - |
|  | **Highest Quality** | - | - | - |
| **Independent**<br>**Validation** | **Strongest Effect Size** | - | - | - |
|  | **Highest Quality** | - | - | - |
| Outcome:<br>MSSS | Predictor: Genes |  |  |  |
| **Training** | **Strongest Effect Size** | 0.16 (20378664, 2, 605) | - | - |
|  | **Highest Quality** | _<sup>1</sup> | - | - |
| **Cross**<br>**Validation** | **Strongest Effect Size** | - | - | 0.58 (31396954, 6, 2) |
|  | **Highest Quality** | - | - | _<sup>1</sup> |
| **Independent**<br>**Validation** | **Strongest Effect Size** | - | 0.06 (31396954, 6, 94) | 0.20 (31396954, 6, 9) |
|  | **Highest Quality** | - | _<sup>1</sup> | _<sup>1</sup> |
| Outcome:<br>ARMSS | Predictor: Genes |  |  |  |
| **Training** | **Strongest Effect Size** | - | - | - |
|  | **Highest Quality** | - | - | - |
| **Cross**<br>**Validation** | **Strongest Effect Size** | - | - | 0.58 (31396954, 6, 2) |
|  | **Highest Quality** | - | - | _<sup>1</sup> |
| **Independent**<br>**Validation** | **Strongest Effect Size** | - | 0.12 (31396954, 6, 94) | 0.17 (31396954, 6, 9) |
|  | **Highest Quality** | - | _<sup>1</sup> | _<sup>1</sup> |

<sup>1</sup>No additional studies available

For modeling MS progression using ordinal EDSS scale (Table 1), we found comparable highest-reported effect sizes between studies that used clinical (i.e., R^2 = 0.67) and MRI (R^2 = 0.64) predictors. The decrease in effect size for best-in class studies was larger for clinical predictors (i.e., R^2 = 0.26) to MRI predictors (R^2 = 0.52). Only MRI predictors reported cross-validation results, which further decreased effect size to R^2 = 0.19. We identified no independent validation cohorts. Blood biomarker predictors achieved much lower effect size in predicting EDSS: the strongest effect size (R^2 = 0.19) was reported by study that included only 23 subjects and achieved technical quality score of 1, whereas highest quality study reported R^2 = 0.06. We identified no cross-validation or independent validation studies for blood predictors of EDSS. Finally, we identified no studies reporting genetic or CSF-biomarker-based predictors of EDSS.

For predicting MS severity (Table 2) measured by MSSS, the strongest reported effect size was R^2 = 0.45 for MRI and R^2 = 0.24 for clinical predictors. However, these were derived from small training cohorts (n=67 for MRI and n=54 for clinical predictors) and were not validated. We identified several studies using genetic predictors of MS severity; the effect sizes for independent validation of MSSS and ARMSS were reported only as correlation coefficients and ranged from Pearson Rho = 0.17-0.2. We did not identify any blood or CSF biomarker-based models of MS severity.

### 3.7 Shiny-App exploration tool

To facilitate independent exploration of the rich dataset we collected beyond Excel worksheet containing all extracted data and deposited as Supplementary Table 1, we also developed Shiny App that allows selective filtering of the data (e.g., to isolate specific predictors, specific outcomes and specific statistical metrics of effect sizes). It can be found at following link: https://jliu159.shinyapps.io/MS_Models_LitSearch_Data_Exploration/ This tool was designed to facilitate comparisons of any future models with the reviewed literature. A user manual can be found in the Supplementary Material.

## 4 Discussion

Technological advances make measuring thousands of genes, transcripts, proteins and metabolites and hundreds of imaging and clinical biomarkers relatively easy and common. Thanks to analogous computational advances these measurements can be aggregated into models expected to elucidate disease mechanisms and provide clinical (e.g., prognostic) value. These are valuable developments; however, to fulfill expectations of providing reproducible knowledge and clinical value, these technological advances must be paired with the rigor of experimental design.

This review shows large potential to improve modeling of clinical disease characteristic in MS. It is startling that 21% of published studies failed to implement *any* of the seven attributes of strong experimental design [9, 11-13, 15] to limit bias and enhance reproducibility. Additional 36% of reviewed studies implemented only one out of seven technical criteria, making this the median attribute of experimental design quality in MS models. This is clearly suboptimal.

This inferior experimental design is compounded by frequent use of small sample sizes (i.e., less than 100 subjects): in fact, for MRI and blood non-genetic biomarker studies, the median cohort sizes were below 100. Considering the complexity of disease mechanisms in polygenic diseases like MS, a modeling cohort of less than 100 MS patients cannot comprise the entire spectrum of disease heterogeneity. Moreover, such small studies are highly susceptible to bias [11, 13], especially when less than 20% used blinding, less than 25% adjusted for covariates, and less than 30% addressed missingness or adjusted the threshold of significance for the number of comparisons performed (sometimes more than hundreds).

Evidence from other scientific areas [10, 11, 13, 14], supported by this paper, shows that poor experimental design, intensified by small cohort sizes, over-estimates effect sizes. This is inevitable, as statistical power is positively associated with cohort- and effect sizes [18]. Consequently, the only way small studies can reach statistical significance is if they demonstrate unusually high effect sizes. These high effect sizes are almost always inflated, as abnormalities in individual transcripts, proteins or metabolites are only mild or moderate, with severe disturbances being incompatible with life [21].

Another under-appreciated aspect of complex modeling algorithms is their incredible power to overfit. Contrary to laymen understanding, it is surprisingly easy to derive seemingly strong models in the training cohorts, especially if one measures comparably higher number of biomarkers to the number of subjects. That is why validation of such models is essential. But validation was included only in 15% of all studies and most of these (56%) used cross-validation rather than independent validation. Indeed, less than 8% of all studies validated their model(s) on completely new set of subjects (i.e., independent validation cohort), which is the gold standard.

Cross-validation (also called rotation estimation or out-of-bag [OOB] testing), re-uses some of the training cohort data by partitioning or resampling the data to train and test models on different iterations. For example, a training cohort may be randomly partitioned (many times) to generate “internal” training and validation splits; this partitioning may be as large as 50:50 split or as small as leaving out only 1 sample. The model then tests accuracy of the predictions of these OOB samples. Because cross-validation does not require any new datasets, it should be included with all studies, not just 10% of them. Although cross-validation is certainly better than no validation, it tends to still overestimate power/accuracy of the classifier in comparison to true independent validation. We have *always* observed decreases in model performance (e.g., predictive accuracy) from training cohort to cross-validation and from cross-validation to independent validation [20-22]. These decreases happen irrespective whether we use clinical data [23], functional data [26], MRI data ([22, 25]), soluble biomarkers ([20, 27]) or genes [23]; and[21], and they are often substantial, especially when comparing cross-validation with true independent validation (e.g., from R^2^ 0.72 in the training cohort to 0.64 in 5-fold cross-validation with 10 repetitions to 0.01 in the independent validation [26]). Please note that the effect sizes for EDSS-based outcomes summarized in Tables 1&2 also show decreasing effect sizes with increasing quality of experimental design, and from training to cross-validation results. Finally, we emphasize that an exceptionally low p-value achieved in the training cohort (even in the cross-validation cohort) does not guarantee the dramatic loss of the model’s accuracy observed in the independent validation cohort [15, 21].

Cross-validation frequently overestimates the accuracy of the model because it often includes a circular argument: somewhere in the modeling process, usually at its very beginning, for example at the feature constriction step, the OOB samples contributed to the construction of the model. These early modeling steps (such as quality control, outlier removal, feature selection), if performed unblinded, introduce bias and are often omitted from the publication altogether (a problem called “selective reporting” [28-30]). Consequently, the introduced bias may not be identified during the review process. Another source of bias that leads to major misinformation in scientific literature is publication bias [33]: where so called “positive” studies (i.e., those that achieved arbitrary p-value <0.05) are published, but “negative” studies, including negative independent validation studies, remain frequently unpublished. This collectively causes unrealistically optimistic view of the reproducibility of the published results.

We initiated this work with the goal of identifying opportunities for advancing modeling of MS outcomes. Based on this work we endorse following recommendations:

1. *Enhance experimental design of future studies*: to minimize bias and maximize reproducibility, no modeling study should fulfill less than four criteria of sound experimental design, and all should include at minimum cross-validation. Studies should also be of sufficient size, inclusive of all MS phenotypes, to increase the probability that the results will be generalizable.
2. *Include most common outcomes (e*.*g*., *EDSS-based) as comparators*. While modeling new, possibly better clinical, or functional outcomes (including PROs) is desirable, unless EDSS-based outcomes are included, it is impossible to compare different models and understand their clinical utility.
3. *Prioritize modeling continuous (or ordinal) over dichotomized outcomes*. Even though EDSS is ordinal scale and EDSS-based severity outcomes (i.e., MSSS and ARMSS) are continuous, 71/138 (51% of) studies used EDSS in dichotomized manner: e.g., predicting progression (yes/no) within certain period. Of the 71 studies that used dichotomized EDSS-based outcomes, dichotomization was not uniform across studies. For example, EDSS worsening was defined as a 1-point increase in one study, 0.5-point increase in other study and increase by 0.5 or 1 points depending on some EDSS threshold, which varied between EDSS 4-6. Without justification for a specific definition of EDSS dichotomization and assurance that this definition was selected before data analyses, non-uniform selection of EDSS-based outcomes may lead to bias, while also preventing comparison between studies. Such call for greater standardization of clinical outcomes has been made previously in MS field [16]. We strongly recommend that even studies that chose to dichotomize EDSS-based outcome include models that predict EDSS as ordinal scale and MSSS/ARMSS as continuous scales. Predicting when and how much progression will occur is a mathematically harder problem than predicting whether a patient might progress. While dichotomized model may predict that two patients will progress over next five years, continuous model may predict that one patient progresses by 3 EDSS points starting next year and other progresses by 0.5 EDSS points at year five. This level of granularity, if validated, provides greater biological insight into mechanisms of disease progression and stronger information gain for clinical management. Because the data (i.e., EDSS) is already collected, applying different modeling strategies, and reporting their outcomes is not difficult.
4. *Report broad and accurate metrics of model’s accuracy*. We observed highly inadequate reporting of metrics of model’s accuracy, at times limited only to p-value. P-values do not reliably reflect model’s accuracy; in fact, one can get a low p-value for aa model that has an inverse relationship with a measured outcome. Or, in large cohorts, a clinically insignificant model, (explaining less than 1% of variance,) may provide a surprisingly low p-value. For continuous outcomes, correlation coefficients only reflect strength of association between measured and predicted outcomes, but not the accuracy of the model: e.g., let’s imagine that measured and predicted outcomes are distributed in perfect (positive) line, resulting in correlation coefficients of 1. However, while the measured EDSS has spread of values between 0 and 10, the predicted EDSS may have different spread of values: e.g. 4 to 6 or 1-2. In fact, such “mis-calibrated” models are quite common. The R^2, reflecting the proportion of the variance explained by the model is preferable to correlation coefficients. But the best indicator of model’s accuracy reflects how closely the model’s predictions match the absolute values of the measured outcomes (i.e., 1:1 line), such as Lin’s concordance coefficient (CCC). Current statistical packages, including freely available options such as R, can calculate all these statistical parameters. Their reporting will provide better assessment of model’s accuracy and would facilitate comparison between studies.
5. *Address clinical utility of the models*. Not all models have, or must have clinical utility; as indicated above, molecular, genetic or cellular biomarker-predictors might be useful by simply linking specific pathophysiological processes or pathways to MS clinical outcomes. However, even these models should assess and publish metrics of clinical utility, such as ROC, accuracy, sensitivity/specificity, and positive and negative predictive values, so that clinicians correctly understand their potential clinical value (or lack of).
6. *Validate most promising observations in the independent cohort(s)*. The low rate of independent validation (i.e., 8% of studies) observed in this meta-analysis is, unfortunately, consistent with similar reports of very low independent validation rates [17]. Because a “lack of validated predictive tools in MS” has been recognized before [18], the funders need to devote more funding to high quality, definite independent validation studies. Analogously, the reviewers and readers should recognize that training cohort data, even cross-validation, has high probability to over-estimate generalizability of the model(s), and reward publications that include independent validation cohorts.
7. *Deposit raw data*. Most journals do not limit the amount of supplementary data. Data sharing is essential for independently validating the algorithms that underlie published models, but also for exploring stronger algorithms/models.

Finally, as evidenced from the summary of current EDSS-based models, we identified great need for developing validated models of MS clinical outcomes using cellular or molecular biomarkers. Vast majority of reviewed models used clinical or MRI predictors. While these may provide clinical value, they are less likely to yield mechanistic insight into MS progression or MS severity, necessary for development of effective treatments for progressive MS or treatments that would abrogate accumulation of disability in patients treated by current disease-modifying agents that successfully limit formation of new lesions.

## Supporting information

Supplemental Shiny App User Manual

## Data Availability

All raw data pertaining to this paper has been deposited as Supplementary information.

https://jliu159.shinyapps.io/MS_Models_LitSearch_Data_Exploration/

## 6 Author Contributions

J. L. and E.K. performed the literature search and extracted all data for this meta-analysis. J.L. analysed the data, generated Figures, Shiny App and contributed to writing of the paper. B.B. construed the project conceptually, guided and supervised all aspects of the study and contributed to the writing of the paper. All authors critically reviewed and edited the manuscript.

## 8 Data Availability Statement

All raw data pertaining to this paper has been deposited as Supplementary information.

