## Supplemental Shiny App User Manual for "Current status and future opportunities in modeling Multiple Sclerosis clinical characteristics"

Shiny App User Guide

Shiny App URL Link: <https://jliu159.shinyapps.io/MS_Models_LitSearch_Data_Exploration/>

This Shiny app was developed with the intention of providing access to the curated database of 302 studies reporting statistical models in the Multiple Sclerosis (MS) field.

**To display data initially**: the user must first select any outcomes of interest and click “refresh.” Several plots of data will load after several seconds.

**Side Panel:** selection criteria to filter to studies of interest.

1. Refresh: select criteria of interest as described below and then click “refresh” button to load up and display several plots of data on the main panel.
2. Metrics: select type of effect size from dropdown menu (All Metrics, p-value, R^2, Pearson R, Spearman Rho, Hazard Ratio, Odds Ratio, AUC, Accuracy, MSE, CCC, negative log likelihood, sensitivity).
3. Clinical Outcomes: selection of clinical outcomes categorized into 3 layers: categories, subcategories, and actual outcomes. Specific keywords can be searched in the search bar. Use checkboxes to select any combination of outcomes of interest.
4. Predictor Type: select any combination of five predictor types: clinical, MRI, genes, blood, and CSF. The latter three are grouped under laboratory biomarkers.
5. Cohort: select combination of cohorts of interest, to look at data from training cohorts, cross-validation cohorts, or independent validation cohorts.

**Main Panel:** click through information in each panel using the tabs at the top of the screen.

1. # Criteria: displays data on the seven experimental design quality criteria. The first plot shows the distribution of studies by number of subjects (y-axis) and number of criteria fulfilled (x-axis). Each datapoint represents a unique study and can be clicked on to be automatically redirected to the PubMed study online.
2. Predictor: displays information on the distribution of studies in each predictor type, as well as the number of subjects per predictor type.
3. Validation: displays how the effect sizes vary between cohorts.
4. Multiple Comparisons: displays how adjusting for multiple comparisons correlates with number of comparisons made in a study as well as the effect sizes reported.
5. Blinding: shows how blinding affects affect size.
6. MS Types: three types of MS subtypes are primarily utilized in MS modeling studies, namely RRMS, SSPMS, and PPMS. The numbers of MS subtypes included in studies (from 0 to all 3) is explored in correlation with effect size.
